# Effect of transitioning virally suppressed children and adolescents with HIV to dolutegravir-based antiretroviral therapy: emulated target trials in a large cohort in South Africa

**DOI:** 10.64898/2026.08.19.26360677

**Authors:** Jennifer A. Brown, Yukteshwar Sookrajh, Luvo Mtila, Noluthando Lushaba, Minenhle Hlabisa, Johan S. van der Molen, Kwena Tlhaku, Mbali Nkosi, Thulani Ngwenya, Thokozani Khubone, Sharana Mahomed, Frédérique Chammartin, Moherndran Archary, Nigel Garrett, Lara Lewis, Jienchi Dorward

## Abstract

**Background:** Global HIV programmes are transitioning virally suppressed children and adolescents with HIV (CAWH) from prior regimens to dolutegravir-based antiretroviral therapy (ART). However, the supporting evidence largely stems from randomised trials in viraemic CAWH. The effect of transition for virally suppressed CAWH is unknown.

**Methods:** We used observational, de-identified data from 724 clinics in KwaZulu-Natal, South Africa. We sequentially emulated three distinct target trials to estimate the effect of transitioning to dolutegravir-based ART in three paediatric populations: i) ages 8-17 years taking efavirenz-based ART, ii) 8-17 years taking ritonavir-boosted lopinavir (LPV/r)-based ART, and iii) 0-7 years taking LPV/r-based ART, all with a last viral load <1,000 copies/mL. The risk difference (RD) of death or viraemia>1,000 copies/mL through 12 and 24 months was estimated using an inverse probability weighting approach.

**Findings:** From January 2020 to August 2024, 37,145 CAWH contributed 454,081 person-trials. In CAWH initially taking efavirenz, the standardised 12-month risk of death or viraemia was 11·9% with continued efavirenz and 6·7% with transition to dolutegravir (RD -5·2 [95% CI -5·8 to -4·6]). In older CAWH initially taking LPV/r, these risks were 17·8% and 9·5%, respectively (RD -8·3 [-10·0 to -6·8]). In younger children, the respective risks were 15·8% and 6·7% (RD -9·0% [-12·7 to -5·4]). Where available, 24-month endpoints showed slightly greater RDs.

**Interpretation:** This large-scale, causal analysis highlights improvements in viral suppression and strongly supports ongoing transition to dolutegravir-based ART for virally suppressed CAWH.

**Funding:** Gates Foundation, National Institute for Health and Care Research, Swiss National Science Foundation

**Research in context:** *Evidence before this study:* We searched PubMed on 23 June 2026 for randomised controlled trials or trial emulations comparing dolutegravir (DTG)-based antiretroviral therapy (ART) with non-DTG-based ART among children and adolescents with HIV (CAWH). The search string is shown in Table S1. Two major trials, ODYSSEY and CHAPAS-4, compared DTG-based with non-DTG-based ART regimens among CAWH with viraemia (i.e. newly starting first-line ART or switching to second-line ART after treatment failure with a non-nucleoside reverse transcriptase inhibitor (NNRTI)-based regimen). Both trials showed better treatment outcomes with DTG-based ART, informing the ongoing programmatic rollout of DTG-based ART for CAWH. In practice, the majority of CAWH eligible for DTG will be virally suppressed on a non-DTG regimen, a group which was not included in ODYSSEY and CHAPAS-4. To date, there is a lack of empirical evidence demonstrating outcomes after paediatric transition to DTG in case of viral suppression. On the one hand, unnecessary ART modification may disrupt adherence; on the other hand, DTG-based ART may be favourable considering the complexity and unpalatability of some prior standard of care regimens. Therefore, there is a need to evaluate the causal effect of transitioning to DTG-based ART in case of viral suppression, specifically among CAWH as a priority group for improving treatment outcomes.

*Added value of this study:* Here, we compared treatment outcomes among three distinct paediatric groups in the South African HIV programme who became eligible to transition to DTG-based ART at different time points: older CAWH initially taking efavirenz (EFV)-based ART, older CAWH initially taking ritonavir-boosted lopinavir (LPV/r)-based ART, and young children initially taking LPV/r-based ART. To our knowledge, this study of 38,041 CAWH is the largest clinical cohort of CAWH receiving DTG and presents the first estimate on the effect of transitioning to DTG-based ART among CAWH with viral suppression. Across all groups, the standardised cumulative risk of death or viraemia was lowered by switching to ART containing DTG compared with remaining on the former standard of care, with risk differences (RDs) ranging from -5·2% to -9·0% at 12 months and increasing over follow-up time.

*Implications of all the available evidence:* We observed substantial improvements in treatment outcomes with transition to DTG. These findings strongly support the continued transition to DTG-based ART among virally suppressed CAWH.

## Introduction

Globally, an estimated 2·4 million children and adolescents up to the age of 19 years are living with HIV^1^. Treatment outcomes in this age group have long lagged behind those observed among adults^2,3^. Before the rollout of dolutegravir (DTG), efavirenz (EFV) was the medication of choice to combine with two nucleoside/nucleotide reverse transcriptase inhibitors (NRTIs) for children and adolescents with HIV (CAWH) aged three years and over initiating antiretroviral therapy (ART), whereas younger children were recommended ritonavir-boosted lopinavir (LPV/r)-based ART due to the risk of

vertically transmitted resistance to EFV. LPV/r-based ART was also used as second-line treatment after failure of a first-line regimen containing a non-nucleoside reverse transcriptase inhibitor (NNRTI) such as EFV.

The ODYSSEY^4,5^ and CHAPAS-4^6^ trials showed superior efficacy of DTG compared with previous standard of care regimens among viraemic CAWH starting first- or second-line ART. Since 2018^7^, DTG-containing ART has been rolled out in several stages, eventually becoming available to children weighing below 20 kg in South Africa in 2023^8^. This programmatic transition included both CAWH newly initiating first- or switching to second-line ART and those already taking ART. However, no randomised controlled trials have assessed the effect of transitioning to DTG-based ART among virally suppressed CAWH, who make up the majority of those eligible for DTG-based ART globally and for whom transition may risk disrupting adherence to established regimens. Considering current guidelines, such trials will no longer be conducted.

Here, we used routinely collected data from the South African HIV programme to estimate the impact of transitioning to DTG-based ART on treatment outcomes among virally suppressed CAWH.

## Methods

### Study design

In the absence of randomised trials, emulated target trials using observational data have been shown to provide valuable insights into the comparative efficacy of different treatments^9,10^. Target trial emulation involves first specifying a causal effect of interest and the hypothetical target trial that would be used to answer it, and then describing how this target trial is emulated using observational data^10^.

We specified three target trials that would estimate the effect of transitioning to DTG-based ART, as compared with remaining on a prior standard of care regimen, among three distinct, virally suppressed paediatric populations with HIV in South Africa: older CAWH (8-17 years) initially taking EFV-based ART, older CAWH initially taking LPV/r-based ART, and younger children (0-7 years) initially taking LPV/r-based ART. We opted for these distinct trial populations to reflect treatment groups who became eligible for DTG-based ART at distinct time points in the South African ART programme. We then emulated these target trials with observational data, using a sequential trials approach to handle the lack of a clear time zero in those not transitioned to DTG^11,12^. The detailed specifications of the target and emulated trials are shown in **Table S2**.

This study is reported according to the Strengthening the Reporting of Observational Studies in Epidemiology (STROBE)^13^ and Transparent Reporting of Observational Studies Emulating a Target Trial (TARGET)^10^ guidelines.

### Setting

South Africa has an adult HIV prevalence of 17%^14^ and is home to 380,000 CAWH aged ≤19 years^1^. Within South Africa, the province of KwaZulu-Natal has the highest HIV prevalence^15^.

Before availability of DTG-based ART, adolescents and children initiating ART while ≥3 years of age would initiate EFV-based ART, with LPV/r-based ART as a second-line regimen in case of treatment failure. Children initiating ART between 4 weeks and <3 years of age started LPV/r-based ART and would largely remain on first-line LPV/r-based ART after reaching 3 years of age. The stepwise rollout of DTG in South Africa is summarised in the **Supplement**.

### Participants

In three trial populations, we included CAWH taking former standard of care treatment from when the DTG rollout started for the respective group until the latest date allowing sufficient follow-up. Accordingly, we included CAWH meeting the following criteria: being 8-17 years old and taking EFV-based ART between January 2020 and August 2023 (allowing 24 months’ follow-up); being 8-17 years old and taking LPV/r-based ART between June 2021 and August 2023 (allowing 24 months’ follow-up); and being 0-7 years old and taking LPV/r-based ART between September 2023 and August 2024 (allowing 12 months’ follow-up as longer follow-up time was infeasible). CAWH were included only in the trial population for which they first became eligible, making these populations mutually exclusive. Across all emulated trials, additional eligibility criteria were: having an ART backbone consisting of lamivudine or emtricitabine (XTC) and either abacavir (ABC), zidovudine (AZT), or, for those aged ≥10 years, tenofovir disoproxil fumarate (TDF); not having had any ART regimen change within the last 180 days; and having a last viral load <1,000 copies/mL taken within the last 12 months.

Children <20 kg were not eligible DTG before availability of dispersible DTG. We did not have weight in the dataset. Therefore, we conservatively set the age cut-off to 8 years, when around 95% of girls and boys are expected to weigh ≥20 kg^16^.

### Outcome measures and exposures

The composite primary endpoint was death or viraemia ≥1,000 copies/mL through 12 (younger LPV/r trial emulation) or 24 months (older EFV and older LPV/r trial emulations). The composite secondary endpoint was death or viraemia ≥50 copies/mL among person-trials with a baseline viral load <50 copies/mL, over the same time periods. The exposure variable was the ART regimen at baseline (i.e., at the time of emulated randomisation) of the respective person-trial, i.e. EFV- or LPV/r-versus DTG-based ART.

### Data sources and data management

We used deidentified data from the South African Three Interlinked Electronic Register database (TIER.Net), a national monitoring system that covers all people receiving ART or tuberculosis treatment in public sector clinics^17,18^. Data are captured by a data clerk after every clinic visit.

For this analysis, we used data from 724 primary care clinics (including 161 mobile clinics) in KwaZulu-Natal, South Africa. We included demographic data, clinical status (including dates of death and transfer-out), ART regimen, laboratory results (CD4 count and viral load), and clinic visit information (visit dates, scheduling, and referral to a decentralised ART programme) of eligible clients. Data were obtained in March 2026 and after review, data closure was set to 31 August 2025 to allow for backlogs in data capture.

### Statistical methods

We used a sequential trials approach to handle the lack of a clear time zero for the start of follow-up and avoid immortal time bias^11,12^. Our study focuses on the estimation of the per protocol causal contrast effect. For each sequential trial initiated at monthly intervals, we assessed eligibility based on updated individual characteristics and assigned participants to the treatment arm compatible with the observed data. Participants can therefore contribute to several person-trials while receiving their former standard of care regimen, but to a maximum of one person-trial on DTG-based ART.

Full statistical details are given in the **Supplement** and **Table S2**. In brief, we fitted weighted pooled logistic regression models for the discrete-time probability of the composite endpoint. Inverse probability of treatment weights (IPTWs) were used to adjust for baseline confounding, and inverse probability of censoring weights (IPCW) were used to account for informative censoring due to loss to follow-up, transfer-out, and regimen change. Stabilised IPTWs were truncated at the first and 99^th^ percentile. Unstabilised IPCWs were truncated at the 99^th^ percentile in the older EFV and older LPV/r trial emulations, and at the 95^th^ percentile in the younger LPV/r trial emulation. IPTW models included the following covariates at baseline of the respective sequential trial: sex, age, municipality, facility type, WHO stage at ART initiation, known history of viraemia, last viral load, time since last viral load, being in a decentralised ART delivery programme (omitted in the younger LPV/r trial emulation), last ART backbone, calendar time, and sequential trial number (see **Table S2** for more information on included covariates). IPCW models included the same baseline covariates and additionally included the following time-varying variables: enrolment in a decentralised ART programme (omitted in the younger LPV/r trial emulation), person-trial follow-up month.

To improve robustness and precision, the weighted outcome model also adjusted for baseline covariates, together with treatment strategy and time variables. We used weighted pooled logistic regression to obtain monthly hazards of the outcome, from which we calculated standardised monthly risks. We obtained 95% confidence intervals by bootstrapping with 500 iterations, sampling with replacement.

We report crude person-trial follow-up, censoring, and treatment outcome information. Standardised risks and 95% CIs are displayed as standardised risk curves and reported at 12 and (where applicable) 24 months with corresponding risk differences (RDs).

An intention-to-treat analysis, which would correspond to the effect of treatment assignment at baseline, was not pursued as transition to DTG was common and expected over time. All analyses were done using the statistical software R version 4.4.0.

## Results

### Participant characteristics

From the overall cohort (**Figure 1**), we included 37,145 CAWH across all three trial emulations. Median age at inclusion in an individual’s first sequential trial was 14 years (IQR 11-16); 20,263 (54·6%) CAWH were female and 16,882 (45·4%) were male.

**Figure 1:**
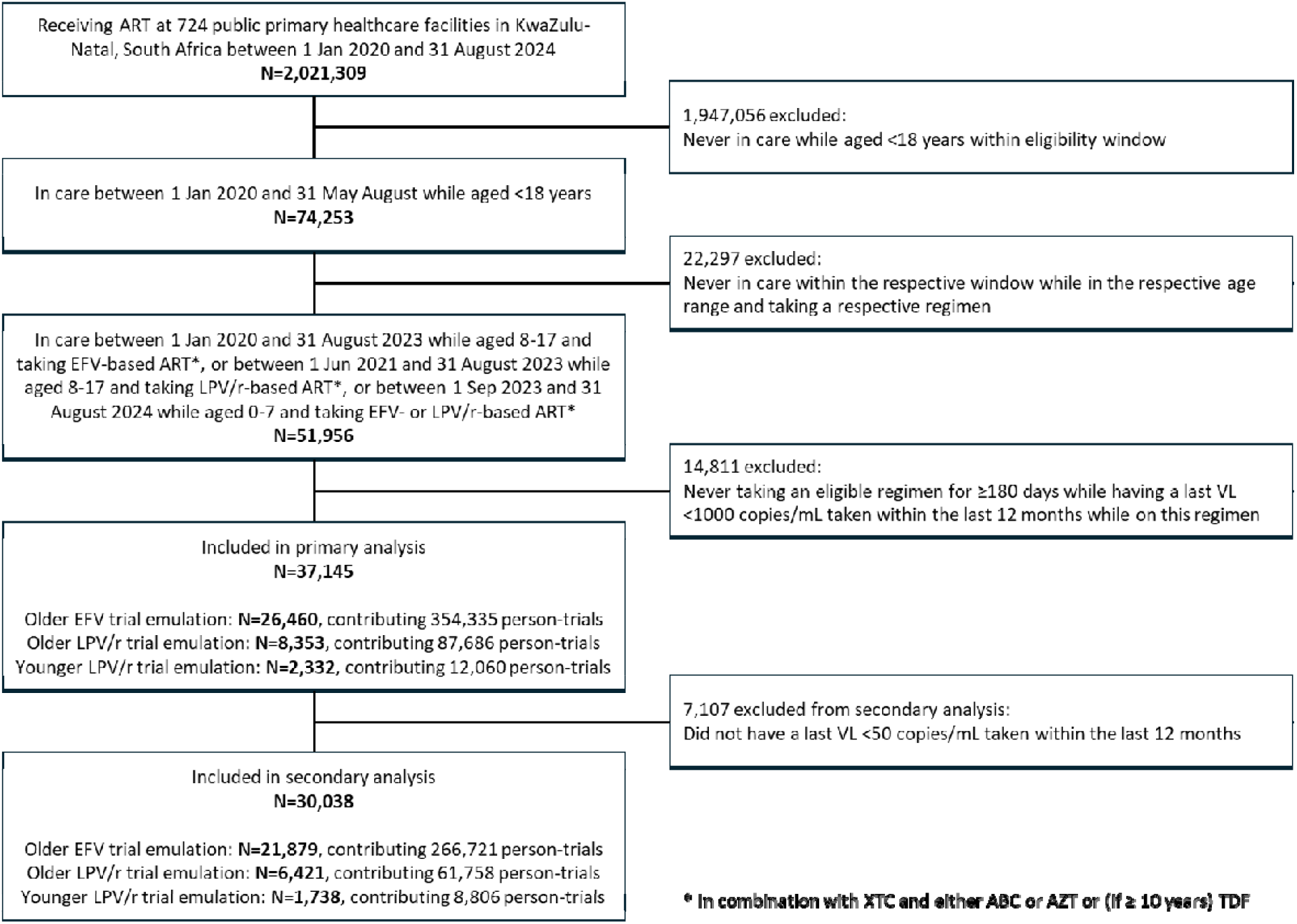
Flow diagram of virally suppressed children and adolescents with HIV eligible to transition to dolutegravir-based antiretroviral therapy in 724 clinics in South Africa. ABC: abacavir; ART: antiretroviral therapy; AZT: zidovudine; DTG: dolutegravir; EFV: efavirenz; LPV/r: ritonavir-boosted lopinavir; TDF: tenofovir disoproxil fumarate; VL: viral load

In the older EFV emulated trial, 26,460 individual CAWH contributed 354,335 person-trials. Of these, 11,583 (3·3%) were taking DTG-based ART; a low proportion of person-trials on DTG-based ART is expected as individuals can contribute to consecutive monthly person-trials while taking a former standard of care regimen but can contribute to maximally one person-trial on DTG. In the older LPV/r emulated trial, we included 8,353 CAWH contributing 87,686 person-trials, of which 2,748 (3·1%) were taking DTG. In the younger LPV/r emulated trial, we included 2,332 CAWH contributing 12,060 person-trials, of which 1,187 (9·8%) were taking DTG. **Table 1** and **Table S3**, respectively, show unweighted and IPTW-weighted person-trial baseline characteristics for the three trial emulations. Treatment groups were balanced after weighting (**Table S3**).

**Table 1:**
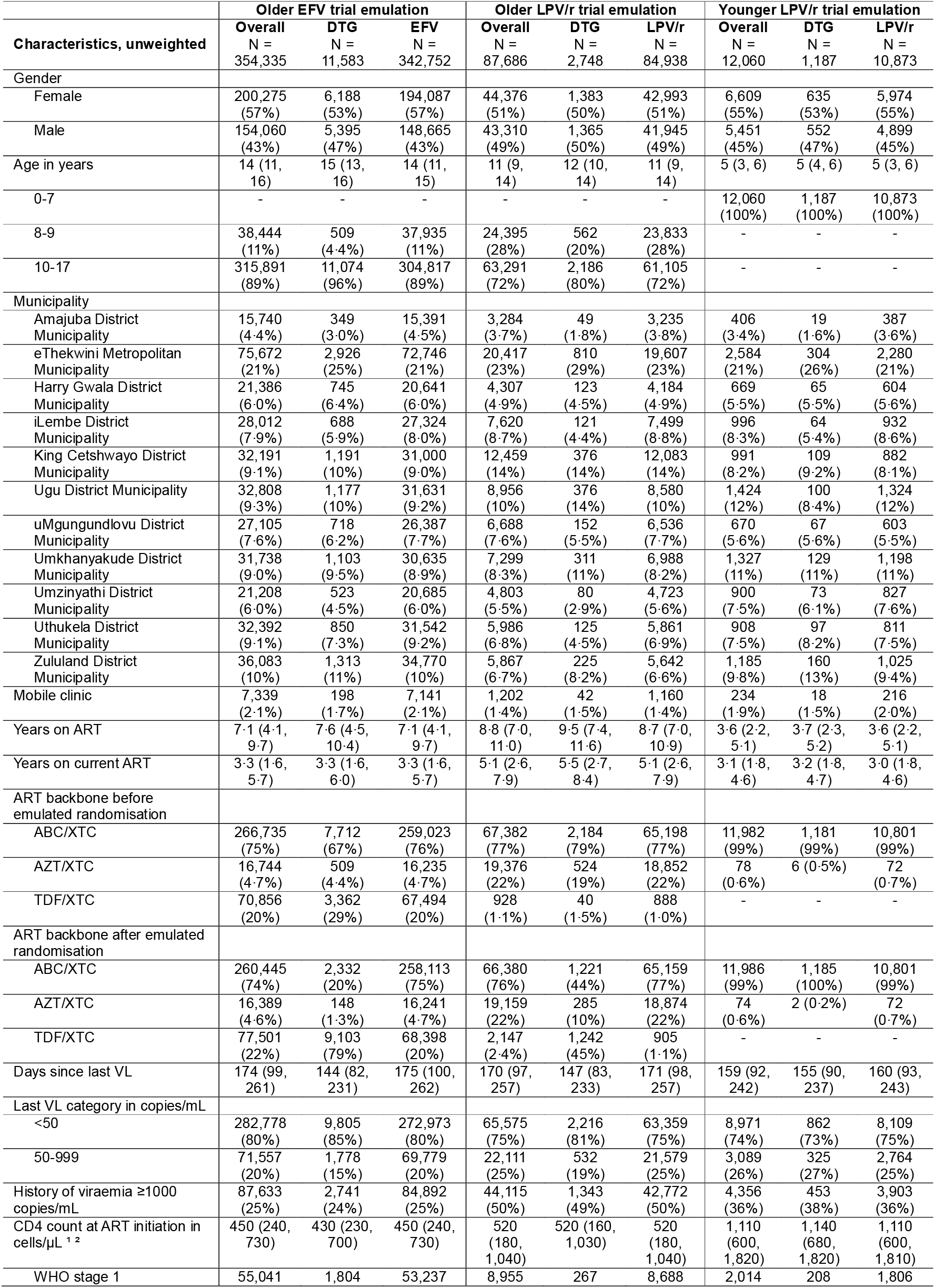
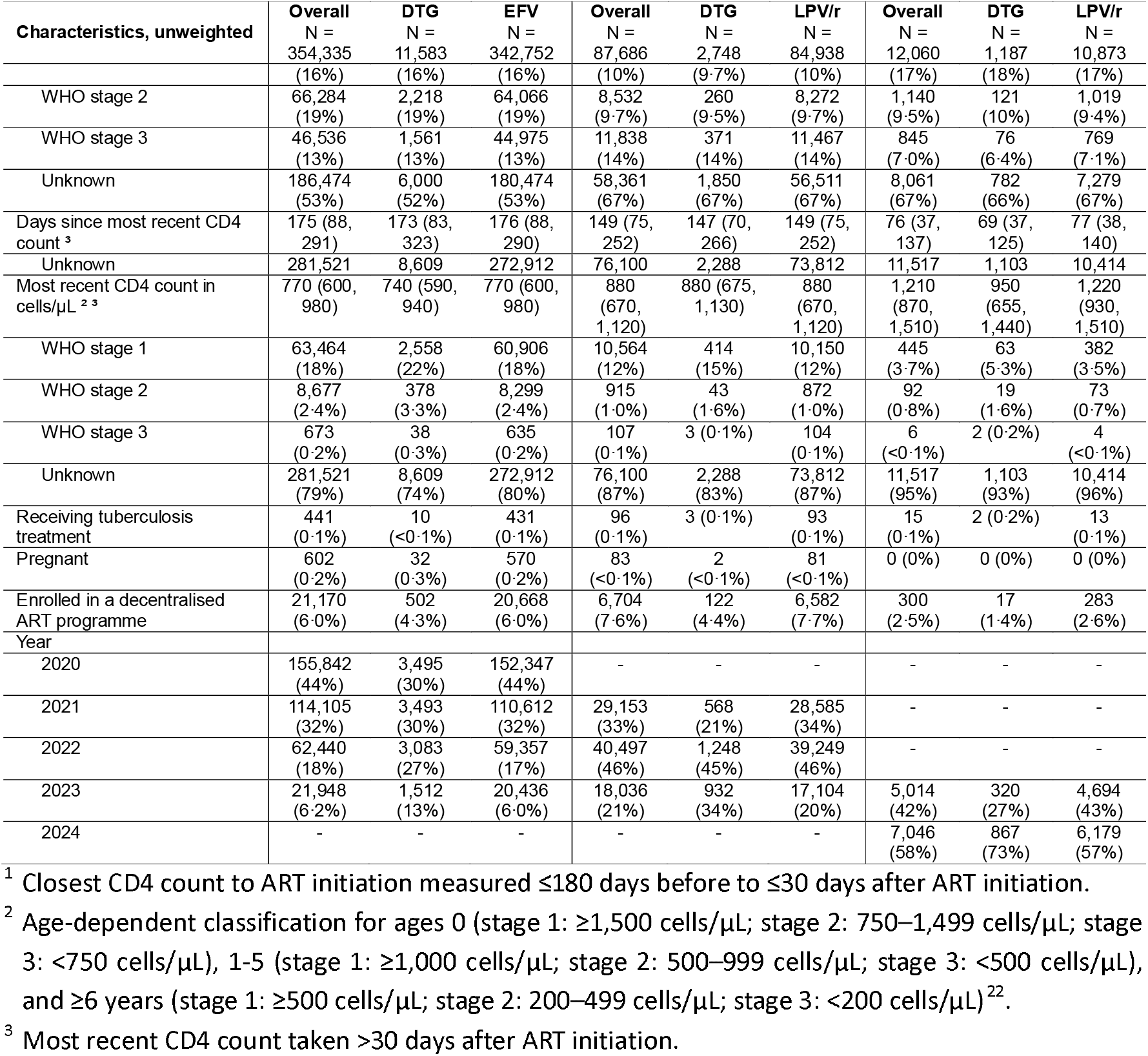
Unweighted characteristics of the pseudo-population at baseline of each person-trial. Categorical variables are indicated as n (%), continuous variables as median (IQR). ABC: abacavir; AZT: zidovudine; DTG: dolutegravir; EFV: efavirenz; IQR: interquartile range; LPV/r: ritonavir-boosted lopinavir; SOC: standard of care; TDF: tenofovir disoproxil fumarate.

### Follow-up and crude outcomes

The older EFV emulated trial encompassed 394,377 person-trial-years of follow-up (20,063 in the DTG group; 374,314 in the EFV group). In the older LPV/r group, there were 85,309 person-trial-years of follow-up (4,652 in the DTG group; 80,657 in the LPV/r group), and in the younger LPV/r group, 6,102 person-trial-years of follow-up (1,043 in the DTG group; 5,059 in the LPV/r group). Crude follow-up, censoring, death, and viraemia outcomes through 12 and 24 months are shown in **Table 2**. Most censoring occurred due to regimen change in the non-DTG groups of each trial emulation as DTG was progressively rolled out.

**Table 2:** Censoring and crude incidence of death and viraemia in CAWH pseudo-populations. Only the first censoring or outcome event is considered. Categorical variables are indicated as n (%), continuous variables as median (IQR). CAWH: children and adolescents with HIV; DTG: dolutegravir; EFV: efavirenz; LPV/r: ritonavir-boosted lopinavir; SOC: standard of care.

|  | Older EFV trial emulation |  |  | Older LPV/r trial emulation |  |  | Younger LPV/r trial emulation |  |  |
| --- | --- | --- | --- | --- | --- | --- | --- | --- | --- |
|  | Overall<br>N =<br>354,335 | DTG<br>N =<br>11,583 | SOC<br>N =<br>342,752 | Overall<br>N =<br>87,686 | DTG<br>N =<br>2,748 | SOC<br>N =<br>84,938 | Overall<br>N =<br>12,060 | DTG<br>N =<br>1,187 | SOC<br>N =<br>10,873 |
| **Follow-up time** |  |  |  |  |  |  |  |  |  |
| Months of follow-up, capped <sup>1</sup> | 13 (6, 22) | 24 (24, 24) | 12 (6, 22) | 10 (5, 18) | 24 (22, 24) | 10 (5, 18) | 5 (3, 10) | 12 (12, 12) | 5 (2, 8) |
| **Crude outcomes through 12 months** |  |  |  |  |  |  |  |  |  |
| Censoring due to loss to follow-up or transfer-out | 15,255 (4.3%) | 503 (4.3%) | 14,752 (4.3%) | 4,108 (4.7%) | 105 (3.8%) | 4,003 (4.7%) | 608 (5.0%) | 84 (7.1%) | 524 (4.8%) |
| Months to censoring due to loss to follow-up or transfer-out | 6 (3, 9) | 7 (4, 9) | 6 (3, 9) | 5 (3, 8) | 6 (4, 10) | 5 (3, 8) | 4 (2, 6) | 7 (5, 9) | 3 (2, 5) |
| Censoring due to regimen change | 126,014 (35.6%) | 507 (4.4%) | 125,507 (36.6%) | 34,549 (39.4%) | 147 (5.3%) | 34,402 (40.5%) | 8,311 (68.9%) | 121 (10.2%) | 8,190 (75.3%) |
| Months to censoring due to regimen change | 6 (3, 9) | 4 (2, 7) | 6 (3, 9) | 6 (3, 9) | 3 (2, 6) | 6 (3, 9) | 4 (2, 7) | 5 (3, 8) | 4 (2, 7) |
| Death | 372 (0.1%) | 8 (0.1%) | 364 (0.1%) | 41 (0.0%) | 1 (0.0%) | 40 (0.0%) | 12 (0.1%) | 3 (0.3%) | 9 (0.1%) |
| Months to death | 6 (3, 9) | 5 (3, 7) | 6 (3, 9) | 5 (3, 9) | 9 (9, 9) | 5 (3, 9) | 8 (5, 9) | 8 (1, 9) | 7 (5, 9) |
| Viraemia | 32,879 (9.3%) | 698 (6.0%) | 32,181 (9.4%) | 12,375 (14.1%) | 216 (7.9%) | 12,159 (14.3%) | 952 (7.9%) | 74 (6.2%) | 878 (8.1%) |
| Months to viraemia | 6 (3, 9) | 7 (5, 10) | 5 (3, 9) | 5 (3, 8) | 6 (4, 9) | 5 (3, 8) | 4 (2, 6) | 5 (2, 7) | 4 (2, 6) |
| **Crude outcomes through 24 months** |  |  |  |  |  |  |  |  |  |
| Censoring due to loss to follow-up or transfer-out | 23,337 (6.6%) | 994 (8.6%) | 22,343 (6.5%) | 5,523 (6.3%) | 208 (7.6%) | 5,315 (6.3%) | - | - | - |
| Months to censoring due to loss to follow-up or transfer-out | 9 (4, 15) | 12 (6, 19) | 9 (4, 15) | 7 (4, 13) | 12 (6, 18) | 7 (4, 12) | - | - | - |
| Censoring due to regimen change | 208,801 (58.9%) | 598 (5.2%) | 208,203 (60.7%) | 55,484 (63.3%) | 168 (6.1%) | 55,316 (65.1%) | - | - | - |
| Months to censoring due to regimen change | 10 (5, 16) | 5 (2, 9) | 10 (5, 16) | 10 (5, 16) | 4 (2, 9) | 10 (5, 16) | - | - | - |
| Death | 581 (0.2%) | 18 (0.2%) | 563 (0.2%) | 69 (0.1%) | 3 (0.1%) | 66 (0.1%) | - | - | - |
| Months to death | 9 (4, 16) | 14 (6, 19) | 9 (4, 15) | 10 (4, 17) | 13 (9, 17) | 10 (4, 17) | - | - | - |
| Viraemia | 46,719 (13.2%) | 1,327 (11.5%) | 45,392 (13.2%) | 15,697 (17.9%) | 367 (13.4%) | 15,330 (18.0%) | - | - | - |
| Months to viraemia | 8 (4, 14) | 12 (7, 18) | 8 (4, 14) | 7 (3, 11) | 10 (5, 18) | 7 (3, 11) | - | - | - |

<sup>1</sup> Within each person-trial, follow-up time was capped at 12 months or 24 months in the younger LPV/r or in the older EFV and older LPV/r trial emulations, respectively. Lower median person-trial follow-up is expected in the non-DTG groups because individuals could be re-enrolled into the non-DTG arm of sequential trials until DTG initiation, resulting in many person-trials ending soon after enrolment due to censoring at treatment change.

### Primary endpoint: standardised risk of death or viraemia ≥1,000 copies/mL

In the older EFV emulated trial, the 12-month risk of death or viraemia ≥1,000 copies/mL was 11·9% (95% CI 11·5 to 12·3) with continued EFV and 6·7% (95% CI 6·2 to 7·2) with transition to DTG (RD - 5·2; 95% CI -5·8 to -4·6) (**Figure 2, Table 3**). The 24-month risk was 21·1% (95% CI 20·2 to 21·9) with continued EFV and 13·5% (95% CI 12·7 to 14·2) with transition to DTG (RD -7·6; 95% CI -8·8 to -6·5).

**Table 3:** Standardised risk of death or viraemia. Risks and absolute risk differences are shown with 95% confidence intervals. DTG: dolutegravir; EFV: efavirenz; IQR: interquartile range; LPV/r: ritonavir-boosted lopinavir; RD: risk difference; SOC: standard of care.

| Standardised cumulative risk | Older EFV trial emulation |  |  | Older LPV/r trial emulation |  |  | Younger LPV/r trial emulation |  |  |
| --- | --- | --- | --- | --- | --- | --- | --- | --- | --- |
|  | DTG | EFV | RD | DTG | LPV/r | RD | DTG | LPV/r | RD |
| **Primary endpoint: death or viraemia $\geq 1,000$ copies/mL** | **N = 11,583** | **N = 342,752** | **N = 354,335** | **N = 2,748** | **N = 84,938** | **N = 87,686** | **N = 1,187** | **N = 10,873** | **N = 12,060** |
| 12 | 6.7% (6.2 to 7.2) | 11.9% (11.5 to 12.3) | -5.2% (-5.8 to -4.6) | 9.5% (8.4 to 10.6) | 17.8% (16.9 to 18.7) | -8.3% (-10.0 to -6.8) | 6.7% (5.2 to 8.2) | 15.8% (12.4 to 19.3) | -9.0% (-12.7 to -5.4) |
| 24 | 13.5% (12.7 to 14.2) | 21.1% (20.2 to 21.9) | -7.6% (-8.8 to -6.5) | 16.0% (14.4 to 17.5) | 27.9% (26.2 to 29.7) | -12.0% (-14.6 to -9.3) | - | - | - |
| **Secondary endpoint: death or viraemia $\geq 50$ copies/mL<sup>1</sup>** | **N = 8,826** | **N = 257,895** | **N = 266,721** | **N = 2,013** | **N = 59,745** | **N = 61,758** | **N = 836** | **N = 7,970** | **N = 8,806** |
| 12 | 15.2% (14.4 to 15.9) | 20.1% (19.5 to 20.7) | -4.9% (-5.9 to -4.0) | 17.9% (16.4 to 19.5) | 28.2% (26.8 to 29.4) | -10.2% (-12.5 to -8.2) | 16.6% (13.9 to 19.3) | 24.3% (19.8 to 28.4) | -7.7% (-13.3 to -2.7) |
| 24 | 30.2% (29.0 to 31.3) | 36.2% (35.1 to 37.3) | -6.1% (-7.7 to -4.3) | 31.1% (28.7 to 33.1) | 44.0% (41.6 to 46.3) | -12.9% (-16.4 to -9.7) | - | - | - |

<sup>1</sup> In the subset of person-trials with a baseline viral load <50 copies/mL.

**Figure 2:**
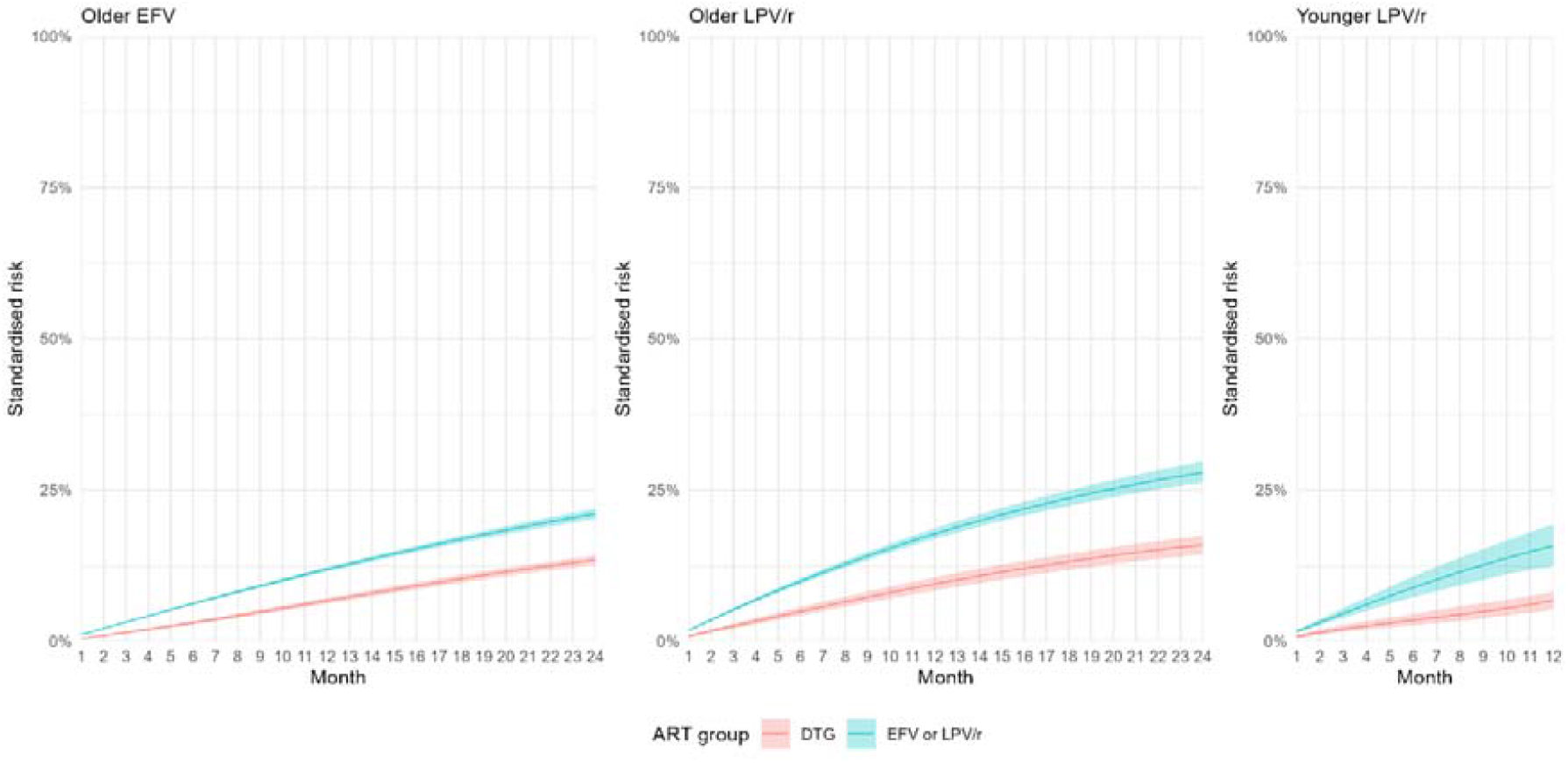
Standardised cumulative risk of death or viraemia ≥1,000 copies/mL. Lines and shading indicate point estimates and 95% confidence intervals, respectively. DTG: dolutegravir; EFV: efavirenz; LPV/r: ritonavir-boosted lopinavir

Within the older LPV/r emulated trial, the 12-month risk was 17·8% (95% CI 16·9 to 18·7) with continued LPV/r and 9·5% (95% CI 8·4 to 10·6) with transition to DTG (RD -8·3, 95% CI -10·0 to -6·8). At 24 months, the standardised risk was 27·9% (95% CI 26·2 to 29·7) and 16·0% (95% CI 14·4 to 17·5), respectively (RD -12·0%; 95% CI -14·6 to -9·3).

Finally, in the younger LPV/r emulated trial, the 12-month risk was 15·8% (95% CI 12·4 to 19·3) with continued LPV/r and 6·7% (95% CI 5·2 to 8·2) with transition to DTG (RD -9·0%; 95% CI -12·7 to -5·4).

### Secondary endpoint: standardised risk of death or viraemia ≥50 copies/mL

For the secondary endpoint of death or viraemia ≥50 copies/mL, analysed among individuals with a last viral load <50 copies/mL at enrolment, overall standardised risks were substantially higher than for the primary endpoint (**Figure S1, Table 3**). Across all emulated trials and time points, the standardised risks were smaller with transition to DTG. In the older EFV emulated trial, the RD at 12 and 24 months was -4·9% (95% CI -5·9 to -4·0) and -6·1% (95% CI -7·7 to -4·3). In the older LPV/r emulated trial, corresponding RDs were -10·2% (95% CI -12·5 to -8·2) and -12·9% (95% CI -16·4 to - 9·7). Finally, in the younger LPV/r emulated trial, the 12-month RD was -7·7% (95% CI -13·3 to -2·7).

## Discussion

In three target trial emulations including 37,145 children and adolescents with HIV who were initially virally suppressed with a non-DTG-containing ART regimen, we estimated that the transition to DTG-based ART substantially reduces incidence of death or viraemia. National and international guidelines now recommend DTG-based ART for almost all CAWH, including transition to DTG for those with viral suppression on a prior standard of care regimen. However, to date there was no clear evidence on the effect of this transition on paediatric treatment outcomes. Here, we report the first causal study quantifying the difference in treatment outcomes with or without transition to DTG in a large cohort of CAWH. Our findings remained consistent regardless of the age group and initial non-DTG ART regimen, or of viral load threshold used to define viraemia, supporting transition to DTG for CAWH with viral suppression on a former standard of care regimen.

In viraemic CAWH, two randomised controlled trials have evaluated virologic treatment outcomes of DTG-based ART. The ODYSSEY trial evaluated non-DTG versus DTG-containing ART in viraemic CAWH who were either ART-naïve or whose first-line ART was failing. Treatment failure (composite of death, clinical, and virological criteria including viraemia ≥400 copies/mL) occurred in 48% (non-DTG) and 31% (DTG) children weighing <14 kg by 96 weeks^5^, and in 22% and 14%, respectively, among CAWH ≥14 kg^4^. The CHAPAS-4 trial (which also included other comparators) among CAWH with viral failure of first-line ART reported viral suppression to <400 copies/mL at 96 weeks for 92.0% of CAWH switched to DTG-versus 80.7% switched to LPV/r-based second-line ART^6^. While there are no randomised trials among virally suppressed CAWH assessing transition to DTG, two studies have been conducted among adults. One, the 2SD randomised trial, showed no effect of transition to DTG among adults with viral suppression taking protease inhibitor-based second-line ART^19^. The other, a trial emulation by our group, estimated that transition from first-line NNRTI-based to first-line DTG-based ART would lead to a small (0·8%) improvement in 12-month viral suppression in adults^20^. In the present study, we found similar reductions in risk of viraemia with DTG to those observed in ODYSSEY and CHAPAS-4. In addition, we are the first to show that the benefits of DTG seen in these clinical trials among CAWH with viraemia extend to those transitioned to DTG while virally suppressed. This difference in findings compared with the adult studies might be caused by greater fluctuation in adherence, and higher incidence of viraemia, in CAWH, allowing for greater effect sizes.

Our study had several strengths. First, this study is, to our knowledge, by far the largest of DTG in CAWH, and further represents one of the larger datasets of CAWH more broadly. This allows for precise estimation of effects in several paediatric populations, including younger children where data are often sparse. Second, they address a previously unanswered question in CAWH transitioning to DTG with viral suppression, who represent the majority of CAWH who will receive DTG globally. Third, the findings reflect routine care outcomes in a resource-limited setting. Fourth, we used robust causal inference methods to emulate target trials in observational data, as no randomised controlled trials are expected to be conducted comparing DTG-based with prior regimens.

The study also had some limitations. First, we were unable to link people between clinics, meaning that CAWH who transferred between included clinics would reflect as separate individuals. Second, previous analyses have shown that viral load data is well-captured in TIER.Net but that death is underreported^18,21^. Third, while we employed causally informed methodology, our findings assume correct model specification and that the available variables allow adequate emulation of randomisation and estimation of per-protocol effects.

The study also has some limitations. First, as with any causal inference analysis based on observational data, our findings rely on assumptions of correct model specification and no unmeasured confounding to adequately emulate randomisation and estimate per-protocol effects. While these assumptions cannot be verified, we believe that our analyses accounted for the most important measured confounders known to influence treatment assignment and outcomes. Second, previous analyses have shown that viral load data is well-captured in TIER.Net but that death is underreported^18,21^. This could lead to an underestimation of our composite endpoint of death or viraemia, though we do not expect under-reporting to differ between treatment strategies and affect our estimated risk difference. Third, we were not able to link people between clinics, meaning that CAWH who transferred between included clinics would reflect as separate individuals.

In conclusion, our findings provide further evidence supporting the benefits of transitionning to dolutegravir-based ART for maintaining viral suppression among children and adolescents with HIV, irrespective of previous regimen, and reinforce current guideline recommendations. While adjusting to the new realities of reduced funding, country HIV programmes should continue to prioritise the rapid rollout of DTG for paediatric care to maximize gains for this vulnerable population.

### Patient consent statement

The cohort study within which this work was conducted was approved by the University of KwaZulu-Natal Biomedical Research Ethics Committee (BE646/17), the eThekwini Municipal Health Department Research Committee, and the KwaZulu-Natal Department of Health Provincial Health Research Ethics Committee (KZ_201807_021), with a waiver of consent for analysis of de-identified, routinely collected data. No emulated target trial protocol was registered. This research was conducted in alignment with the Declaration of Helsinki.

## Data Availability

We cannot publicly share the data used for this analysis because of the legal and ethical requirements regarding the use of routinely collected clinical data in South Africa. Interested parties can request access to the data from the KwaZulu-Natal Provincial Department of Health, the eThekwini Municipality Health Unit and the South African National Department of Health TB/HIV Information System (contact details obtainable upon request to JD).

## Author contributions

Conceptualisation: JD. Funding acquisition: JD, NG, JAB. Methodology: JD, JAB, LL, FC, MA. Formal analysis: JAB, JD, LL, FC, MN. Data curation: YS, LM, NL, TN, MH, TK, JvdM. Project administration: JD, NG, KT, JvdM, YS, SM. Writing – original draft: JAB. Writing – review and editing: all.

## Acknowledgements

This work was supported, in whole or in part, by the Gates Foundation (INV-073793). The conclusions and opinions expressed in this work are those of the authors alone and shall not be attributed to the Foundation. Under the grant conditions of the Foundation, a Creative Commons Attribution 4.0 License has already been assigned to the Author Accepted Manuscript version that might arise from this submission. JAB is funded by the Swiss National Science Foundation (P500PM_221966 and P5R5-3_239083, to JAB). JD is funded by the National Institute for Health and Care Research (NIHR, CL-2022–13–005, to JD) for this research project. The views expressed in this publication are those of the authors and not necessarily those of the NIHR, the National Health Service, or the UK Department of Health and Social Care. The funders had no role in the design, conduct, or reporting of this study.

We thank the eThekwini Municipality, the Umkhanyakude District Municipality, and uMgungundlovu District Municipality Health Units, the KwaZulu-Natal Department of Health, and the staff and patients at the participating healthcare facilities.

