## Supplement for "Effect of transitioning virally suppressed children and adolescents with HIV to dolutegravir-based antiretroviral therapy: emulated target trials in a large cohort in South Africa"

**Methods – setting: DTG rollout in South Africa**

In South Africa, DTG was rolled out in several stages. Guidelines published in late 2019 introduced DTG-based ART for adults, adolescents, and children weighing ≥20 kg^1^. For people already receiving EFV-based first-line ART, transition to DTG-based first-line ART initially necessitated recent viral suppression (<50 copies/mL or twice 50-999 copies/mL) within the last 6 months; these criteria were subsequently loosened. After reviewing the data, we set the start of the DTG rollout for older CAWH established on EFV-based ART to January 2020. The 2019 guidelines also introduced DTG in second-line ART after confirmed virological failure. However, they did not provide guidance on transition to DTG-based ART among people who were virally suppressed with LPV/r-based first- or second-line ART. Upon review of the data, we determined June 2021 as the start of the paediatric transition from LPV/r- to DTG-based ART. 2023 national guidelines then introduced DTG-based ART for CAWH ≥4 weeks of age and ≥3 kg^2^. Transition within first-line ART was now possible for CAWH taking EFV-based ART or taking LPV/r-based ART for less than two years, regardless of viral load result. CAWH taking LPV/r-based ART for more than two years were eligible to transition if they had a viral load <1,000 copies/mL within the last 12 months. In practice, dispersible DTG (required for children weighing <20 kg) became more widely available from around September 2023, which we consequently set as the start date for eligibility in the younger LPV/r trial emulation.

**Methods – statistical methods: weighting and outcome models**

IPTWs were calculated from propensity scores which we obtained from pooled logistic regression in person-trial baseline data for the outcome of ART treatment (i.e., DTG or non-DTG). Covariates were baseline variables of the respective sequential trial, specifically: sex, age, municipality, facility type, WHO stage at ART initiation, known history of viraemia, last viral load, time since last viral load, being in a decentralised ART delivery programme (omitted in the younger LPV/r trial emulation), last ART backbone, calendar time, and sequential trial number (see **Table S2** for more information on included covariates).

Within each sequential trial, person-trials were censored at loss to follow-up (midpoint between last attended and missed next scheduled visit), transfer out of the clinic (we were unable to link people between clinics), or change of the ART anchor drug (guideline-conform NRTI backbone changes were permitted). We computed unstabilised inverse probability of censoring weights (IPCWs) for loss to follow-up and transfer-out, and separately for regimen change. For this, we fitted pooled logistic regressions using expanded datasets (with each person-trial having one row per follow-up month), with loss to follow-up or transfer-out (combined) or regimen change in the next month as the outcome. Covariates were as outlined above, with the addition of the time-varying variables of current enrolment in a decentralised ART programme (omitted in the younger LPV/r trial emulation), and person-trial follow-up month (**Table S2**). With these models, we predicted monthly hazards of being censored, from which we calculated cumulative monthly risk of remaining uncensored and corresponding unstabilised IPCWs. Stabilised IPTWs were truncated at the first and 99^th^ percentile. Unstabilised IPCWs were truncated at the 99^th^ percentile in the older EFV and older LPV/r trial emulation, and at the 95^th^ percentile in the younger LPV/r trial emulation.

Finally, we fitted logistic regression models using expanded datasets, with the endpoint occurring in the subsequent month as the outcome, treatment as the exposure variable of interest, and all above-mentioned baseline and time variables as covariates, applying IPTWs and IPCWs. We then predicted monthly hazards through 12 (younger LPV/r trial emulation) or 24 (older EFV and older LPV/r trial emulation) months under the assumption that no CAWH were transitioned to DTG, or that all CAWH were transitioned to DTG. We calculated cumulative monthly risks from the cumulative monthly hazards within each person-trial, then calculated standardised monthly risks as the mean cumulative monthly risk across person-trials.

**Table S1: Search strategy in PubMed.**

| **Concept** | **Description** |
| --- | --- |
| Population | Child OR adolescent OR teen OR paediatric OR pediatric |
| Treatment | dolutegravir |
| Outcome | (viral load) OR (viral suppression) |
| Study design | randomised OR randomized OR emulated OR emulation |

**Table S2: Specification of the target and emulated trials.** ART: antiretroviral therapy; ABC: abacavir; AZT: zidovudine; DTG: dolutegravir; EFV: efavirenz; LPV/r: ritonavir-boosted lopinavir; TDF: tenofovir disoproxil fumarate; XTC: lamivudine or emtricitabine.

| **Protocol** | **Target trials** | **Emulated trials** |
| --- | --- | --- |
| Eligibility criteria | *Older EFV trial:*  Being in care in a participating clinic between 1 January 2020 and 31 August 2023 while aged 8-17 years old and while taking EFV-based ART.  *Older LPV/r trial:*  Being in care in a participating clinic between 1 June 2021 and 31 August 2023 while aged 8-17 years old and while taking LPV/r-based ART.  *Younger LPV/r trial:*  Being in care in a participating clinic between 1 September 2023 and 31 August 2024 while aged 0-7 years and while taking LPV/r-based ART. | Same. Participants are included only in the trial population for which they first became eligible, making these populations mutually exclusive. |
|  | ART backbone consisting of ABC, AZT, or (if aged ≥10 years) TDF together with XTC. | Same. |
|  | No ART regimen change in the last 180 days. | Same. |
|  | Last viral load result <1,000 copies/mL. | Same |
|  | Last viral load result measured within 12 months before enrolment and while on the current regimen. | Same. |
| Treatment strategies | *Older EFV trial:*  Remain on EFV-based ART.  *Older LPV/r trial:*  Remain on LPV/r-based ART.  *Younger LPV/r trial:*  Remain on LPV/r-based ART. | Same. |
|  | Switch to DTG-based ART, consisting of DTG together with XTC and either ABC, AZT, or (if aged ≥10 years) TDF. | Same. |
| Assignment procedures | Participants are randomly assigned and are aware of the strategy they are assigned to (open-label) | Treatment assignment is assumed to be conditionally random given the measured baseline covariates^a^ included in the inverse probability of treatment weighting model. |
| Follow-up | *Older EFV trial:*  24 months, starting at randomisation.  *Older LPV/r trial:*  24 months, starting at randomisation.  *Younger LPV/r trial:*  12 months, starting at randomisation. | Same, starting at emulated randomisation of the person-trial. |
| Outcomes: primary endpoint | Incidence of death or viraemia ≥1,000 copies/mL. | Same. |
| Outcomes: secondary endpoints | Incidence of death or viraemia ≥50 copies/mL among participants with a last viral load <50 copies/mL at enrolment. | Same. |
| Causal contrast | Per-protocol analysis. Participants are censored upon transfer-out, loss to follow-up, or regimen change (backbone changes are permitted so long as the backbone continues to consist of either ABC, AZT, or if aged ≥10 years, TDF, together with XTC). | Observational analogue thereof. |
| Identifying assumptions | Inverse probability of censoring weighting adequately estimates the outcomes that would have been observed had censored individuals remained in care receiving their assigned treatment. | Same. Additionally, inverse probability of treatment weighting adequately emulates randomisation. |
|  | Incident measured viraemia is an acceptable proxy for incident (measured or unmeasured) viraemia. | Same. |
| Data analysis plan | The primary and secondary endpoints are assessed through pooled logistic regression weighted for inverse probability of remaining uncensored (weights derived using baseline^a^ and time-varying^b^ covariates). | Same with additional inverse probability of treatment weighting as described above to emulate randomisation. |

^a^	Categorical baseline covariates: sex (male or female), municipality (11 categories), facility type (mobile clinic or not), CD4 stage at ART initiation (with age-dependent categorisation^3^ and a missing category), known history of viraemia ≥1,000 copies/mL (binary), last known viral load category (<50 copies/mL or 50-999 copies/mL; omitted for the secondary endpoint where a last VL <50 copies/mL is required), being enrolled in a decentralised ART programme with ART pickup outside the facility (binary; omitted for the younger LPV/r trial emulation due to ineligibility of those aged <5 years), last ART backbone (ABC/XTC, AZT/XTC, or TDF/XTC). Continuous baseline covariates: age in years and age squared, days since last viral load measurement, calendar time (defined as days since start of enrolment for the respective target trial) with cubic splines, and sequential trial number (sequential trial 1 includes all individuals at their earliest eligibility which may be after the start of enrolment for the respective trial emulation, differentiating this variable from calendar time).

^b^	Categorical time-varying covariates: current enrolment in a decentralised ART programme with ART pickup outside the facility (binary; omitted for the younger LPV/r trial emulation due to ineligibility of those aged <5 years). Continuous time-varying covariates: person-trial follow-up month with cubic splines.

**Table S3: Weighted characteristics of the pseudo-population at baseline of each person-trial.** Categorical variables are indicated as n (%), continuous variables as median (IQR). ABC: abacavir; AZT: zidovudine; DTG: dolutegravir; EFV: efavirenz; IQR: interquartile range; LPV/r: ritonavir-boosted lopinavir; TDF: tenofovir disoproxil fumarate.

|  | **Older EFV trial emulation** | | | **Older LPV/r trial emulation** | | | **Younger LPV/r trial emulation** | | |
| --- | --- | --- | --- | --- | --- | --- | --- | --- | --- |
| **Characteristics, weighted** | **Overall**  N = 351,820 | **DTG**  N = 9,077 | **EFV**  N = 342,743 | **Overall**  N = 87,219 | **DTG**  N = 2,283 | **LPV/r**  N = 84,936 | **Overall**  N = 12,002 | **DTG**  N = 1,129 | **LPV/r**  N = 10,873 |
| Gender |  |  |  |  |  |  |  |  |  |
| Female | 198,680 (56%) | 4,951 (55%) | 193,729 (57%) | 44,133 (51%) | 1,148 (50%) | 42,985 (51%) | 6,564 (55%) | 607 (54%) | 5,957 (55%) |
| Male | 153,140 (44%) | 4,126 (45%) | 149,014 (43%) | 43,086 (49%) | 1,136 (50%) | 41,951 (49%) | 5,438 (45%) | 522 (46%) | 4,916 (45%) |
| Age in years | 14 (11, 16) | 14 (12, 16) | 14 (11, 16) | 11 (9, 14) | 12 (10, 14) | 11 (9, 14) | 5 (3, 6) | 5 (4, 6) | 5 (3, 6) |
| 0-7 | - | - | - | - | - | - | 12,002 (100%) | 1,129 (100%) | 10,873 (100%) |
| 8-9 | 37,745 (11%) | 547 (6·0%) | 37,198 (11%) | 24,163 (28%) | 513 (22%) | 23,650 (28%) | 0 (0%) | 0 (0%) | 0 (0%) |
| 10-17 | 314,075 (89%) | 8,531 (94%) | 305,545 (89%) | 63,056 (72%) | 1,770 (78%) | 61,286 (72%) | - | - | - |
| Municipality |  |  |  |  |  |  |  |  |  |
| Amajuba District Municipality | 15,558 (4·4%) | 332 (3·7%) | 15,226 (4·4%) | 3,233 (3·7%) | 52 (2·3%) | 3,181 (3·7%) | 394 (3·3%) | 28 (2·5%) | 366 (3·4%) |
| eThekwini Metropolitan Municipality | 75,315 (21%) | 2,124 (23%) | 73,191 (21%) | 20,381 (23%) | 607 (27%) | 19,773 (23%) | 2,581 (22%) | 252 (22%) | 2,329 (21%) |
| Harry Gwala District Municipality | 21,286 (6·1%) | 596 (6·6%) | 20,690 (6·0%) | 4,291 (4·9%) | 118 (5·2%) | 4,173 (4·9%) | 684 (5·7%) | 79 (7·0%) | 605 (5·6%) |
| iLembe District Municipality | 27,700 (7·9%) | 604 (6·7%) | 27,096 (7·9%) | 7,510 (8·6%) | 129 (5·6%) | 7,381 (8·7%) | 981 (8·2%) | 82 (7·3%) | 898 (8·3%) |
| King Cetshwayo District Municipality | 31,999 (9·1%) | 865 (9·5%) | 31,134 (9·1%) | 12,401 (14%) | 332 (15%) | 12,069 (14%) | 999 (8·3%) | 104 (9·2%) | 895 (8·2%) |
| Ugu District Municipality | 32,597 (9·3%) | 868 (9·6%) | 31,730 (9·3%) | 8,954 (10%) | 279 (12%) | 8,675 (10%) | 1,406 (12%) | 122 (11%) | 1,284 (12%) |
| uMgungundlovu District Municipality | 26,850 (7·6%) | 630 (6·9%) | 26,220 (7·7%) | 6,629 (7·6%) | 151 (6·6%) | 6,479 (7·6%) | 659 (5·5%) | 56 (5·0%) | 603 (5·5%) |
| Umkhanyakude District Municipality | 31,552 (9·0%) | 850 (9·4%) | 30,702 (9·0%) | 7,307 (8·4%) | 236 (10%) | 7,071 (8·3%) | 1,332 (11%) | 135 (12%) | 1,197 (11%) |
| Umzinyathi District Municipality | 20,998 (6·0%) | 481 (5·3%) | 20,518 (6·0%) | 4,735 (5·4%) | 82 (3·6%) | 4,653 (5·5%) | 886 (7·4%) | 75 (6·7%) | 811 (7·5%) |
| Uthukela District Municipality | 32,077 (9·1%) | 743 (8·2%) | 31,334 (9·1%) | 5,918 (6·8%) | 120 (5·3%) | 5,798 (6·8%) | 894 (7·5%) | 78 (6·9%) | 817 (7·5%) |
| Zululand District Municipality | 35,886 (10%) | 984 (11%) | 34,901 (10%) | 5,859 (6·7%) | 176 (7·7%) | 5,683 (6·7%) | 1,186 (9·9%) | 118 (10%) | 1,068 (9·8%) |
| Mobile clinic | 7,270 (2·1%) | 170 (1·9%) | 7,100 (2·1%) | 1,200 (1·4%) | 35 (1·5%) | 1,164 (1·4%) | 231 (1·9%) | 20 (1·8%) | 211 (1·9%) |
| Years on ART | 7·1 (4·1, 9·7) | 7·5 (4·4, 10·3) | 7·1 (4·1, 9·7) | 8·8 (7·0, 11·0) | 9·3 (7·4, 11·5) | 8·8 (7·0, 11·0) | 3·6 (2·2, 5·1) | 3·8 (2·3, 5·2) | 3·6 (2·2, 5·1) |
| Years on current ART | 3·3 (1·6, 5·7) | 3·4 (1·7, 6·0) | 3·3 (1·6, 5·7) | 5·1 (2·6, 7·9) | 5·4 (2·7, 8·2) | 5·1 (2·6, 7·9) | 3·1 (1·8, 4·6) | 3·2 (1·8, 4·7) | 3·0 (1·8, 4·6) |
| ART backbone before emulated randomisation |  |  |  |  |  |  |  |  |  |
| ABC/XTC | 264,419 (75%) | 6,397 (70%) | 258,023 (75%) | 67,069 (77%) | 1,800 (79%) | 65,269 (77%) | 11,924 (99%) | 1,122 (99%) | 10,802 (99%) |
| AZT/XTC | 16,617 (4·7%) | 420 (4·6%) | 16,197 (4·7%) | 19,221 (22%) | 453 (20%) | 18,768 (22%) | 77 (0·6%) | 7 (0·6%) | 70 (0·6%) |
| TDF/XTC | 70,784 (20%) | 2,260 (25%) | 68,523 (20%) | 930 (1·1%) | 31 (1·4%) | 898 (1·1%) | - | - | - |
| ART backbone after emulated randomisation |  |  |  |  |  |  |  |  |  |
| ABC/XTC | 259,208 (74%) | 2,081 (23%) | 257,127 (75%) | 66,279 (76%) | 1,049 (46%) | 65,229 (77%) | 11,930 (99%) | 1,127 (100%) | 10,802 (99%) |
| AZT/XTC | 16,322 (4·6%) | 118 (1·3%) | 16,204 (4·7%) | 19,036 (22%) | 245 (11%) | 18,791 (22%) | 72 (0·6%) | 1 (0·1%) | 71 (0·6%) |
| TDF/XTC | 76,290 (22%) | 6,878 (76%) | 69,412 (20%) | 1,904 (2·2%) | 989 (43%) | 916 (1·1%) | - | - | - |
| Days since last VL | 174 (99, 260) | 159 (89, 246) | 174 (99, 261) | 169 (97, 256) | 157 (87, 242) | 170 (97, 257) | 159 (92, 242) | 157 (92, 235) | 159 (92, 242) |
| Last VL category in copies/mL |  |  |  |  |  |  |  |  |  |
| <50 | 281,029 (80%) | 7,505 (83%) | 273,524 (80%) | 65,313 (75%) | 1,795 (79%) | 63,518 (75%) | 8,929 (74%) | 841 (74%) | 8,088 (74%) |
| 50-999 | 70,791 (20%) | 1,572 (17%) | 69,219 (20%) | 21,907 (25%) | 489 (21%) | 21,418 (25%) | 3,072 (26%) | 288 (26%) | 2,784 (26%) |
| History of viraemia ≥1000 copies/mL | 86,957 (25%) | 2,188 (24%) | 84,769 (25%) | 43,866 (50%) | 1,134 (50%) | 42,732 (50%) | 4,352 (36%) | 424 (38%) | 3,928 (36%) |
| CD4 count at ART initiation in cells/µL ¹ ² | 450 (240, 730) | 440 (230, 710) | 450 (240, 730) | 520 (180, 1,040) | 550 (180, 1,040) | 520 (180, 1,040) | 1,110 (600, 1,810) | 1,090 (610, 1,810) | 1,110 (600, 1,810) |
| WHO stage 1 | 54,632 (16%) | 1,393 (15%) | 53,238 (16%) | 8,903 (10%) | 229 (10%) | 8,675 (10%) | 2,006 (17%) | 190 (17%) | 1,816 (17%) |
| WHO stage 2 | 65,844 (19%) | 1,728 (19%) | 64,116 (19%) | 8,485 (9·7%) | 220 (9·6%) | 8,265 (9·7%) | 1,141 (9·5%) | 112 (10·0%) | 1,028 (9·5%) |
| WHO stage 3 | 46,220 (13%) | 1,208 (13%) | 45,012 (13%) | 11,774 (13%) | 308 (13%) | 11,466 (14%) | 839 (7·0%) | 77 (6·9%) | 762 (7·0%) |
| Unknown | 185,124 (53%) | 4,748 (52%) | 180,376 (53%) | 58,057 (67%) | 1,527 (67%) | 56,530 (67%) | 8,016 (67%) | 749 (66%) | 7,267 (67%) |
| Days since most recent CD4 count ³ | 176 (88, 291) | 174 (83, 309) | 176 (88, 290) | 149 (75, 252) | 142 (66, 262) | 149 (75, 252) | 78 (39, 139) | 65 (37, 117) | 80 (39, 144) |
| Unknown | 279,298 | 6,853 | 272,446 | 75,657 | 1,921 | 73,735 | 11,459 | 1,067 | 10,392 |
| Most recent CD4 count in cells/µL ² ³ | 770 (600, 980) | 750 (590, 950) | 770 (600, 980) | 880 (670, 1,120) | 870 (680, 1,130) | 880 (670, 1,120) | 1,210 (880, 1,510) | 950 (640, 1,440) | 1,230 (950, 1,510) |
| WHO stage 1 | 63,185 (18%) | 1,918 (21%) | 61,267 (18%) | 10,544 (12%) | 326 (14%) | 10,217 (12%) | 448 (3·7%) | 46 (4·1%) | 401 (3·7%) |
| WHO stage 2 | 8,665 (2·5%) | 280 (3·1%) | 8,385 (2·4%) | 911 (1·0%) | 33 (1·4%) | 878 (1·0%) | 90 (0·7%) | 14 (1·3%) | 75 (0·7%) |
| WHO stage 3 | 671 (0·2%) | 27 (0·3%) | 645 (0·2%) | 108 (0·1%) | 3 (0·1%) | 105 (0·1%) | 5 (<0·1%) | 1 (0·1%) | 4 (<0·1%) |
| Unknown | 279,298 (79%) | 6,853 (75%) | 272,446 (79%) | 75,657 (87%) | 1,921 (84%) | 73,735 (87%) | 11,459 (95%) | 1,067 (95%) | 10,392 (96%) |
| Receiving tuberculosis treatment | 440 (0·1%) | 6 (<0·1%) | 433 (0·1%) | 95 (0·1%) | 2 (0·1%) | 93 (0·1%) | 14 (0·1%) | 1 (<0·1%) | 13 (0·1%) |
| Pregnant | 596 (0·2%) | 24 (0·3%) | 573 (0·2%) | 83 (<0·1%) | 2 (<0·1%) | 81 (<0·1%) | 0 (0%) | 0 (0%) | 0 (0%) |
| Enrolled in a decentralised ART programme | 20,949 (6·0%) | 469 (5·2%) | 20,479 (6·0%) | 6,618 (7·6%) | 124 (5·4%) | 6,494 (7·6%) | 298 (2·5%) | 16 (1·4%) | 283 (2·6%) |
| Year |  |  |  |  |  |  |  |  |  |
| 2020 | 153,822 (44%) | 3,071 (34%) | 150,751 (44%) | - | - | - | - | - | - |
| 2021 | 113,146 (32%) | 3,025 (33%) | 110,121 (32%) | 28,806 (33%) | 580 (25%) | 28,226 (33%) | - | - | - |
| 2022 | 62,691 (18%) | 2,054 (23%) | 60,637 (18%) | 40,288 (46%) | 1,080 (47%) | 39,207 (46%) | - | - | - |
| 2023 | 22,160 (6·3%) | 927 (10%) | 21,233 (6·2%) | 18,126 (21%) | 623 (27%) | 17,503 (21%) | 4,970 (41%) | 456 (40%) | 4,514 (42%) |
| 2024 | - | - | - | - | - | - | 7,031 (59%) | 673 (60%) | 6,359 (58%) |

^1^	Closest CD4 count to ART initiation measured ≤180 days before to ≤30 days after ART initiation

^2^	Age-dependent classification for ages 0 (stage 1: ≥1,500 cells/µL; stage 2: 750–1,499 cells/µL; stage 3: <750 cells/µL), 1-5 (stage 1: ≥1,000 cells/µL; stage 2: 500–999 cells/µL; stage 3: <500 cells/µL), and ≥6 years (stage 1: ≥500 cells/µL; stage 2: 200–499 cells/µL; stage 3: <200 cells/µL)^3^

^3^	Most recent CD4 count taken >30 days after ART initiation

**Figure S1: Standardised cumulative risk of death or viraemia ≥50 copies/mL.** Lines and shading indicate point estimates and 95% confidence intervals, respectively. DTG: dolutegravir; EFV: efavirenz; LPV/r: ritonavir-boosted lopinavir

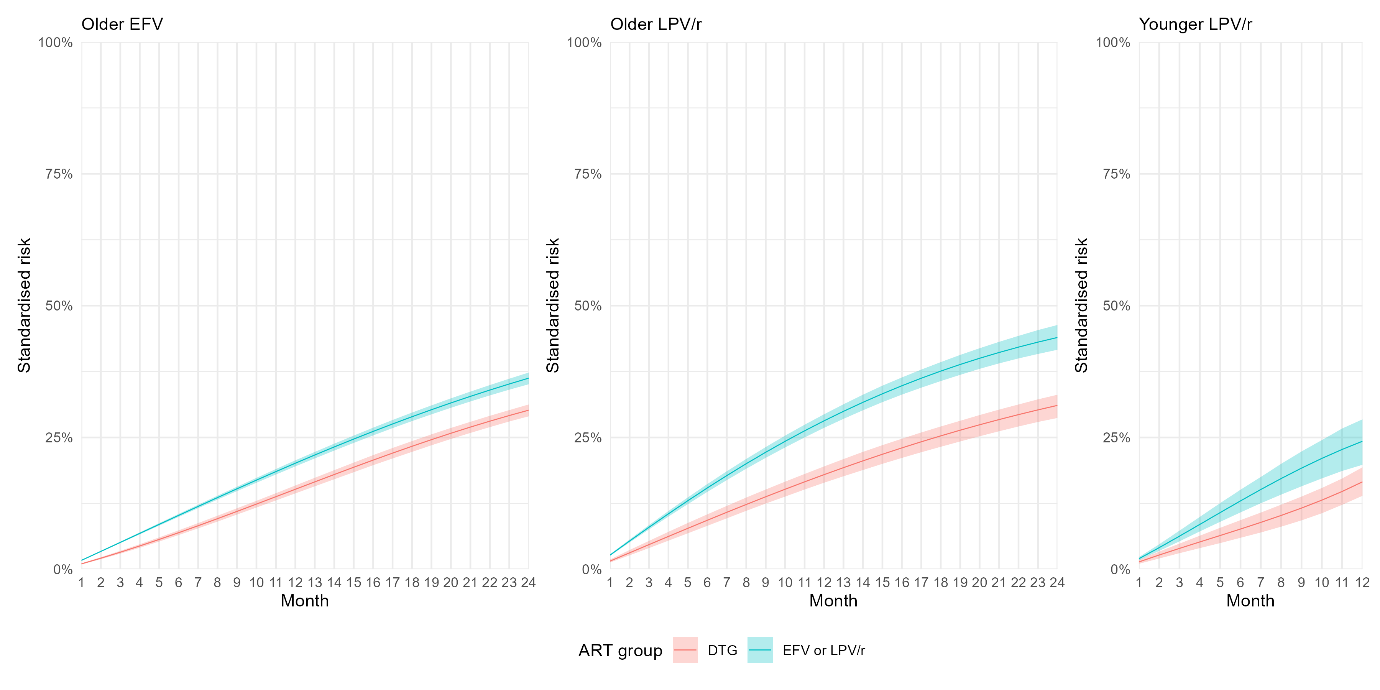
